# Mental health conditions and incident cancer: a prospective cohort study of 402,255 UK Biobank participants

**DOI:** 10.1101/2025.05.23.25328170

**Authors:** Mohammed Sherif Amin, Frederick K Ho, Solange Parra-Soto, Ziyi Zhou, Shinya Nakada, Ike Dhiah Rochmawati, Carlos Celis-Morales, Jill P Pell

## Abstract

1

**Background:** Mental health conditions (MHCs) affect both psychological health and biological systems and have been linked to cancer risk. However, evidence from epidemiological studies remains inconsistent.

**Aim:** The study aims to investigate the associations of five MHCs (depressive disorders (DD), anxiety disorders (AD), bipolar disorder (BD), schizophrenia (SZ), and post-traumatic stress disorder (PTSD)) with overall and site-specific cancer risk. Dose-response relationship by depressive symptom severity and effect modifications by sex were also explored.

**Methods:** A population-based prospective cohort study was conducted on 402,255 UK Biobank participants, who were free from cancer, with a median follow-up of 13.4 years. Cox proportional hazard models were used to examine the associations of the five MHCs, and self-reported depressive symptom severity with overall and site-specific incident cancers, adjusting for sociodemographic, lifestyle and health-related confounders.

**Results:** Over a median follow-up of 13.4 years, 68,065 (17%) incident cancer cases were recorded. DD (HR 1.18; 95% CI 1.13-1.23), AD (HR: 1.17, 95%CI: 1.11-1.24), and BD (HR: 1.29, 95%CI: 1.11-1.51) were associated with increased overall cancer risk. No significant association was found for SZ and PTSD. The associations of DD (HR: 1.27, 95%CI: 1.18-1.35) and BD (HR: 1.54, 95%CI: 1.26-1.88) were only significant in men. AD and DD were positively associated with lung, blood, and liver cancers, while AD was also associated with prostate cancer. A dose-response relationship was observed between depressive symptom severity and cancer risk.

**Conclusion:** MHCs were associated with a higher risk of overall and some site-specific cancers. While causality cannot be assumed, diagnoses of MHCs could be useful for cancer risk stratification and prevention.

## 2 Introduction

The health burden from cancer is rising worldwide, with 20 million cases reported in 2022 [1]. Europe, in particular, faces a disproportionate impact, with the region contributing 22.4% of the global cancer cases while representing only 10% of the population [2]. Consistently, cancer incidence in the UK has risen by 0.8% annually over the past 25 years [3] despite the significant advancement in preventive initiatives targeting well-established risk factors such as smoking [4] and alcohol consumption [5]. Notably, these factors only contribute to about 37% of cancer cases [4], highlighting the need to explore other prevalent modifiable risk factors to mitigate the escalating burden.

Mental health conditions (MHCs) have been previously linked to various physical conditions [6], especially cancer [7]. Traditionally, studies examined MHCs as a consequence of cancer. However, since Blumberg et al.’s [8] pioneering study in 1954 suggested a bidirectional relation, numerous studies have investigated the link between MHCs and cancer risk.

Nevertheless, epidemiological evidence regarding this association is largely inconsistent [9]. Some studies reported a higher cancer risk [10–12], while others demonstrated either non-significant associations [13] or even a lower risk for certain cancers [6,13]. Although population-based differences in key risk factors may contribute to this inconsistency, methodological limitations are likely more influential [9]. For instance, the reliance on self-reported data by most studies [10–12,14,15] introduces recall bias and affects results validity. A study comparing participants’ ability to recall their depression symptoms [16] found that less than half of the participants recalled their key symptoms, and recall ability largely depends on the condition severity, a factor that is often overlooked by most studies.

Additionally, most studies [12,15,17,18] had a small sample size, restricting the investigations to a single or pair of MHCs and limiting their statistical power to investigate site-specific cancers. A further limiting factor was the inadequate adjustment for confounding factors [9]. While crucial behavioural risk factors like smoking, alcohol consumption, physical activity levels, and co-morbidities such as diabetes and hypertension can significantly influence both MHCs [19,20] and cancer development [21–23], the majority of studies [6,12,17,24] did not account for these variables.

The current study aimed to address these limitations by investigating the associations between five common MHCs (depressive disorders (DD), anxiety disorders (AD), bipolar disorder (BD), schizophrenia (SZ), and post-traumatic stress disorder (PTSD)) with overall and site-specific cancer incidence using the UK Biobank (UKB). Additionally, the dose-response relationship by depressive symptom severity and effect modifications by sex was also explored.

## 3 Methods

### 3.1 Study Design

The UKB is a population-based cohort that recruited 502,493 participants between 2006 and 2010 across England, Wales, and Scotland [25]. Participants provided comprehensive data through self-administered touch screen questionnaires and in-person assessments, covering various demographics, behavioural habits, medical history, and physical measurements [26]. Health outcomes were tracked through linkage with UK administrative data, including hospital records and death registries [26]. A detailed description of the UKB is available at https://www.ukbiobank.ac.uk.

### 3.2 Mental Health Conditions

MHCs were ascertained through a confirmed diagnosis of DD, AD, BD, SZ, and PTSD identified through linked inpatient hospital admission records. Records were available until March 2021 for data from England and Scotland, while Wales’s data were available till February 2018. In the UKB, condition diagnosis was coded using ICD-10; therefore, participants diagnosed under the following codes were included in the study: F32/F33 for DD, F40-F43 for AD, F31 for BD, F20 for SZ, and F43.1 for PTSD.

Participants diagnosed with each disorder at or before the baseline assessment (between 2006 and 2010) were classified as "diagnosed" to that specific condition, with the rest of the UKB participants serving as the comparison group. Diagnoses made after the baseline but before any cancer diagnosis were also classified as “diagnosed”, after which the follow-up started. Figure 1 shows a flow diagram of the selection process.

**Figure 1.**
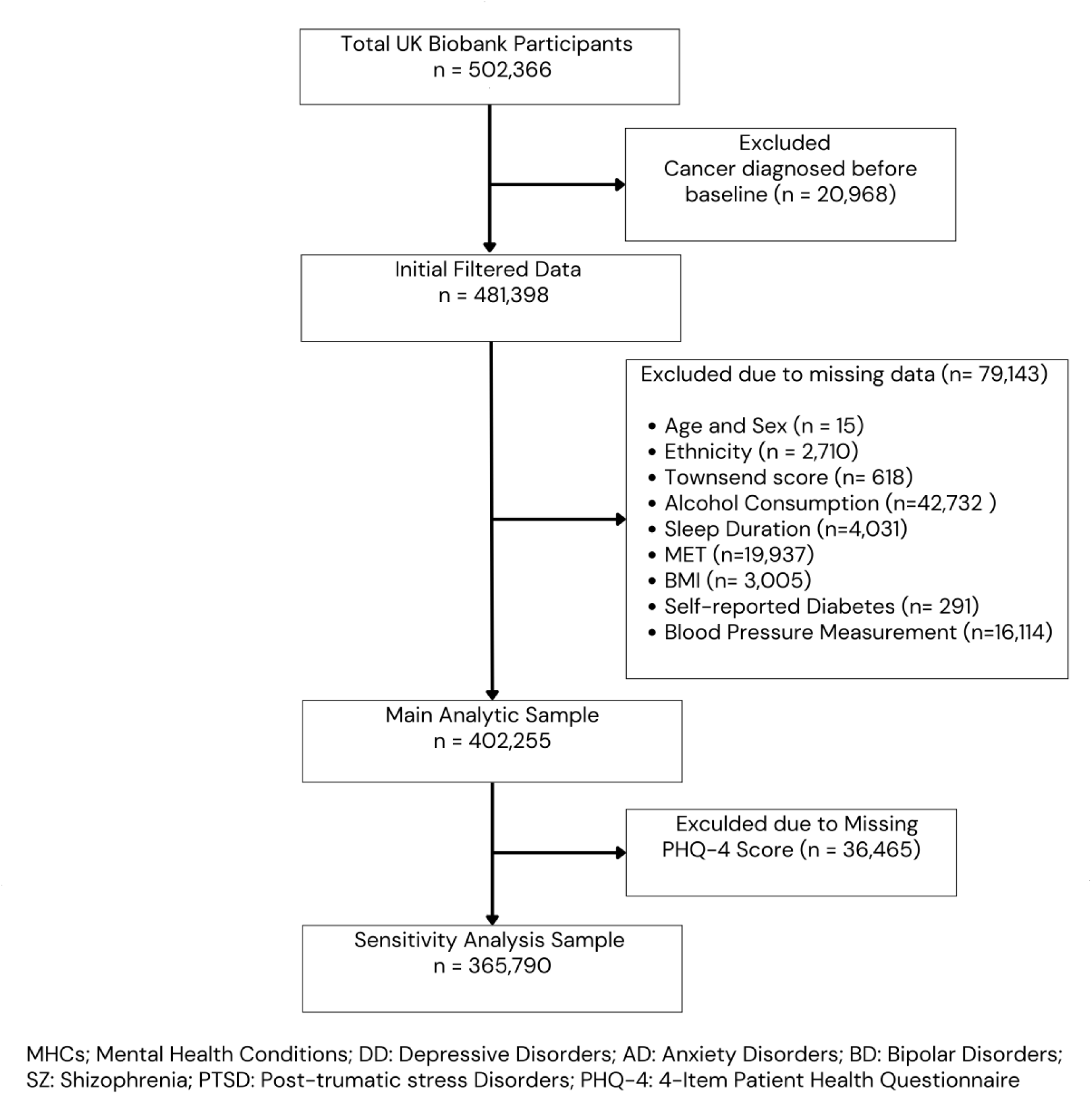
Flow Diagram of the study sample selection.

DD and AD symptoms severity were assessed using the 4-item Patient Health Questionnaire (PHQ-4) collected at baseline. Each question is scored from 0 ("not at all") to 3 ("nearly every day"), and the total score ranges from 0 (no symptoms) to 12 (severe symptoms) [27].

### 3.3 Cancer outcomes

To maintain a cancer-free baseline, those with a pre-baseline cancer diagnosis (n=20,968) were excluded. Overall and site-specific incident cancers were identified as the first eligible code in either inpatient and day-case hospital records or death records.

Accordingly, the "overall cancer" variable represented the post-baseline incident cases under ICD-10 codes C00-C39, C43, and C50-C97 (i.e., all cancers except non-melanotic skin and bone cancers). For site-specific analyses, the study analysed eight common types of cancer: breast (C50), ovarian (C56), uterine (C54-C55), prostate (C61), lung (C33-C34), blood (C81-C96), colorectal (C18-C20), and liver (C22).

### 3.4 Covariates

Covariates were selected based on prior evidence of potential confounding or mediating effects [9], including sociodemographic, behavioural/lifestyle, and health-related factors retrieved from the UKB’s baseline assessment.

#### 3.4.1 Sociodemographic Factors

Age, sex, and ethnicity were self-reported. Townsend area deprivation index was obtained from the postcode of residence and is derived using aggregated data on unemployment, car and home ownership, and household overcrowding [28].

#### 3.4.2 Behavioural/Lifestyle Factors

Smoking was self-reported as "Current", "Previous", or "Never", with any other category recoded as missing in the study. Alcohol consumption was self-reported as units per week (1 unit = 10 ml of ethanol). Physical activity was self-reported using the International Physical Activity Questionnaire Short Form as total Metabolic Equivalent Task (MET). Sleep duration was self-reported based on hours of sleep over a 24-hour period. Lastly, processed meat consumption was self-reported as the average intake over the past year, coded as "Never", "Less than once a week", "Once a week", "2-5 times a week", 5-6 times a week", "once or more daily" [29].

#### 3.4.3 Health-Related Factors

Body Mass Index (BMI) was calculated based on participants’ weight and height (kilograms per meter squared (kg/m^2^)). Diabetes was self-reported based on a physician’s diagnosis as "yes" or "no". Systolic Blood Pressure (SBP) was measured either using an automated device or a sphygmomanometer and reported in millimetres of mercury (mmHg). Lastly, female menopausal status ("Yes" or "No") was considered, as hormonal changes during menopause can influence mood disorders like DD and AD [30] and increase the risk of certain cancers, such as breast and uterine cancer [31].

### 3.5 Ethical approval

The study is conducted under UKB’s generic approval (Ref: 11/NW/0382) and indexed in the UKB under project number 71392.

### 3.6 Statistical analysis

Descriptive statistics established baseline characteristics, stratified by sex and MHC diagnosis, using means (standard deviation [SDs]) for normally distributed continuous variables, medians (Interquartile Range [IQR]) for skewed data, and frequencies and percentages for categorical variables.

The association between MHCs and overall and site-specific cancer incidence was examined using Cox proportional hazard models, producing Hazard Ratios (HR) and corresponding 95% Confidence Intervals (95%CI). Participants with missing data for included covariates were excluded from the analysis (n=79,143) (Figure 1). The dependent variable was the time until cancer occurrence, calculated as the years from the first MHC diagnosis to the cancer diagnosis, death, or study end, whichever came first. All analyses were conducted using a one-year landmark to reduce the potential of reverse causality.

Three incrementally adjusted Cox models were used. Model 1 adjusted for sociodemographic factors, model 2 additionally adjusted for lifestyle/behavioural factors, and model 3 further included health-related factors. Model 3 for female-specific cancers (e.g., breast, ovarian, uterine) was also adjusted for menopausal status. Proportional hazard (PH) assumptions were assessed using Schoenfeld residuals. Initially, models were conducted for each MHC and overall cancer, followed by site-specific cancers. Following previous literature [32], site-specific cancers were only analysed if there was a minimum of five cases per predictor variable to ensure reliability.

Interaction terms were included in the fully adjusted models (model 3) to examine the sex-moderating effect, with further sex stratification for conditions showing significant interactions. A sensitivity analysis using standardised PHQ-4 scores was conducted to assess the impact of symptom severity on the association. All analyses were performed using R version 4.3.1 and set at a significance level of alpha = 0.05.

## 4 Results

Of the 402,255 UK Biobank participants, 39,304 (9.8%) MHCs cases were identified (DD: 25,616 [6.4%], AD: 21,892 [5.4%], BD: 1,510 [0.4%], SZ: 1,719 [0.4%], PTSD: 477 [0.1%]). Over a median follow-up of 13.4 (IQR: 12.6, 14.3) years, 68,065 (17%) incident cancer cases were observed. Site-specific cancer incidence is detailed in Supplementary Table 1

Overall, participants were mostly of white ethnicity (95%) and generally healthy in terms of lifestyle factors, with moderate alcohol consumption (11 units/week) and over half (55%) reporting never smoking (Table 1). Diabetes prevalence was low (4.5%). The mean SBP was elevated (138 mmHg), placing participants within the ‘elevated blood pressure’ category according to the European Society of Cardiology (ESC) guidelines [33].

**Table 1.** Baseline Characteristics of Included Participants (n = 402,255) Stratified by MHC Diagnosis.

| Characteristic | Overall<br>N = 402,255 | Undiagnosed<br>N = 362,951 | Diagnosed<br>N = 39,304 | p-value<sup>4</sup> |
| --- | --- | --- | --- | --- |
| Age<sup>1</sup> (years) | 56 (8) | 56 (8) | 57 (8) | <0.001 |
| sex<sup>2</sup> |  |  |  | <0.001 |
| Female | 214,015 (53%) | 189,493 (52%) | 24,522 (62%) |  |
| Male | 188,240 (47%) | 173,458 (48%) | 14,782 (38%) |  |
| Ethnicity<sup>2</sup> |  |  |  | <0.001 |
| White | 380,847 (95%) | 343,419 (95%) | 37,428 (95%) |  |
| Black | 6,235 (1.6%) | 5,714 (1.6%) | 521 (1.3%) |  |
| Chinese | 1,277 (0.3%) | 1,226 (0.3%) | 51 (0.1%) |  |
| Mixed | 2,360 (0.6%) | 2,062 (0.6%) | 298 (0.8%) |  |
| South Asian | 7,975 (2.0%) | 7,314 (2.0%) | 661 (1.7%) |  |
| Any other | 3,561 (0.9%) | 3,216 (0.9%) | 345 (0.9%) |  |
| Deprivation<sup>1</sup> | -1.37 (3.04) | -1.45 (3.00) | -0.69 (3.34) | <0.001 |
| Smoking Status<sup>2</sup> |  |  |  | <0.001 |
| Current | 41,452 (10%) | 34,910 (9.6%) | 6,542 (17%) |  |
| Never | 219,829 (55%) | 201,336 (55%) | 18,493 (47%) |  |
| Previous | 140,974 (35%) | 126,705 (35%) | 14,269 (36%) |  |
| Alcohol<sup>3</sup> (units/week) | 11 (3, 23) | 11 (3, 23) | 8 (2, 21) | <0.001 |
| Sleep Duration<sup>1</sup> (hour) | 7.16 (1.08) | 7.15 (1.04) | 7.17 (1.37) | 0.004 |
| Total physical activity (MET-min/week)<sup>1</sup> | 2,422 (2,439) | 2,438 (2,432) | 2,281 (2,501) | <0.001 |
| Processed Meat Consumption<sup>1</sup> |  |  |  | <0.001 |
| Never | 37,547 (9.3%) | 33,384 (9.2%) | 4,163 (11%) |  |
| Less than once a week | 121,657 (30%) | 110,004 (30%) | 11,653 (30%) |  |
| Once a week | 117,335 (29%) | 106,214 (29%) | 11,121 (28%) |  |
| 2-5 times a week | 109,667 (27%) | 99,012 (27%) | 10,655 (27%) |  |
| 5-6 times a week | 12,787 (3.2%) | 11,468 (3.2%) | 1,319 (3.4%) |  |
| once or more daily | 3,262 (0.8%) | 2,869 (0.8%) | 393 (1.0%) |  |
| BMI<sup>1</sup> (kg/m<sup>2</sup>) | 27.3 (4.7) | 27.2 (4.6) | 28.3 (5.5) | <0.001 |
| Self-reported Diabetes<sup>2</sup> | 18,006 (4.5%) | 15,341 (4.2%) | 2,665 (6.8%) | <0.001 |
| SBP (mmHg) | 138 (19) | 138 (19) | 137 (19) | <0.001 |
| Menopausal Status<sup>2</sup> |  |  |  | <0.001 |
| No | 52,377 (13%) | 47,260 (13%) | 5,117 (13%) |  |
| Yes | 128,630 (32%) | 114,092 (31%) | 14,538 (37%) |  |
| Not applicable/ unknown | 221,248 (55%) | 201,599 (56%) | 19,649 (50%) |  |
| Follow-up Time<sup>3</sup> (year) | 13.4 (12.6, 14.3) | 13.4 (12.6, 14.3) | 13.2 (11.9, 14.1) | <0.001 |

<sup>1</sup>Mean, (SD); <sup>2</sup>n (%); <sup>3</sup>Median (IQR)
<sup>4</sup>Welch Two Sample t-test; Pearson's Chi-squared test; Wilcoxon rank sum test
Significance level set at alpha = 0.05
MET: Metabolic Equivalent Task; BMI: Body Mass Index; SBP: Systolic Blood Pressure

Table 1 presents baseline characteristics stratified by MHC status. Participants with MHCs exhibited poorer health profiles compared to those without MHCs, including higher smoking (diagnosed: 17%, undiagnosed: 9.6%) and diabetes prevalence (diagnosed: 6.8%, undiagnosed: 4.2%) and lower MET scores. Notably, in the sex-stratified characteristics (Supplementary Table 2), men demonstrated higher mean BMI and SBP, and smoking, alcohol consumption, and diabetes rates than women.

Associations between MHCs and overall cancer incidence are presented in Figure 2. In Model 1, three of the five MHCs investigated were associated with overall cancer risk: DD (HR: 1.18, 95%CI: 1.13-1.23), AD (HR: 1.17, 95%CI: 1.11-1.24) and BD (HR: 1.29, 95%CI: 1.11-1.51). Although slightly attenuated, these associations remained statistically significant in Model 3, with HRs of 1.13 (1.08 - 1.18) for DD, 1.15 (1.09 - 1.21) for AD, and 1.22 (1.05 - 1.43) for BD. Meanwhile, no significant association was found for SZ and PTSD across the three models. The PH assumption was met for all models except for the DD fully adjusted model (p-value=0.01).

**Figure 2.**
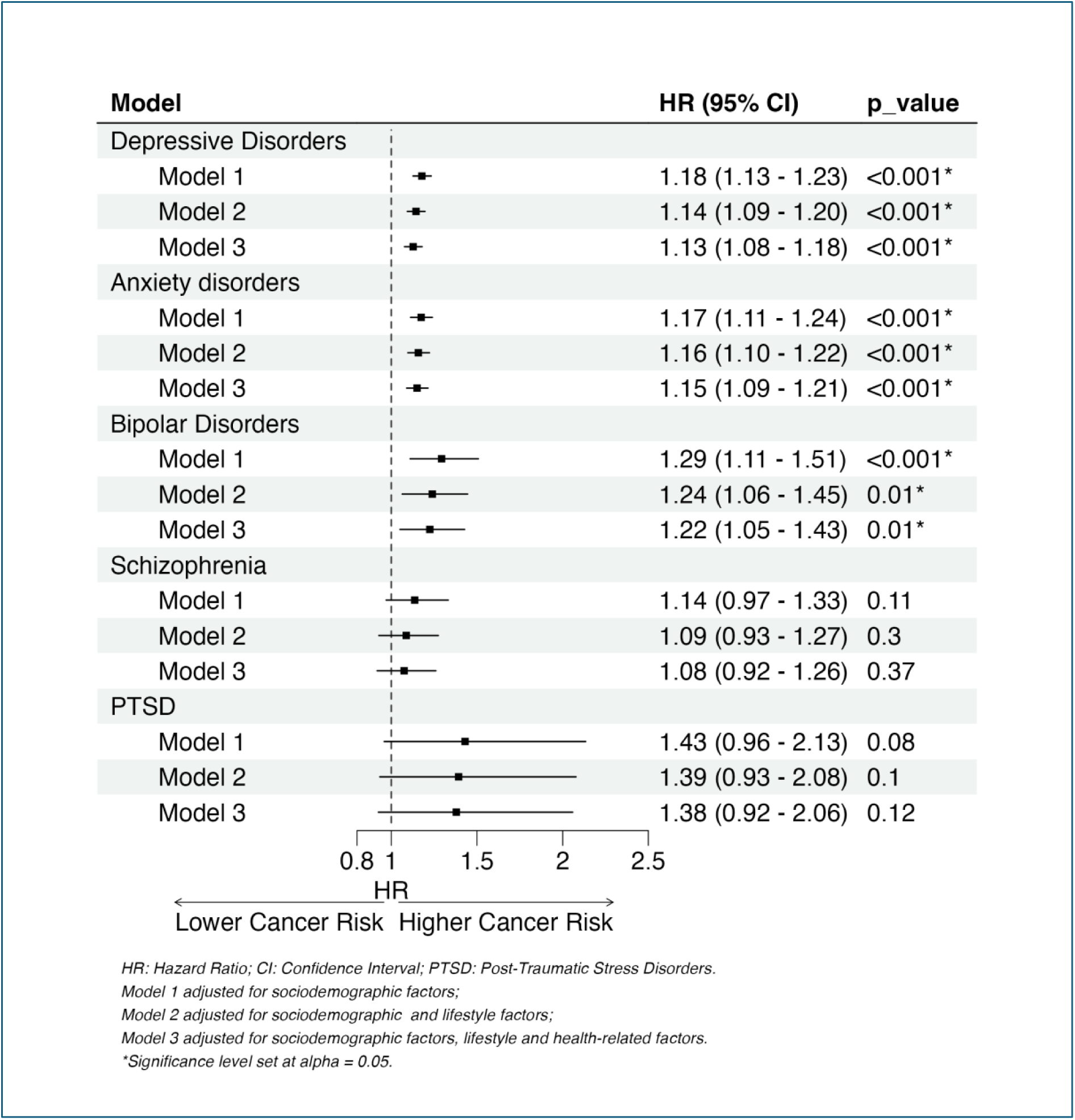
Forest Plot of the Associations between MHCs and Overall Cancer Incidence (n=402,255)

Interaction terms between each MHC and sex were tested using likelihood ratio tests. Only the interactions for DD and BD were statistically significant (Supplementary Table 3), thus both were further stratified by sex. The sex-stratified analysis only found significant associations of DD (HR: 1.27, 95%CI: 1.18-1.35) and BD (HR: 1.54, 95%CI: 1.26-1.88) in men (Supplementary Figure 1).

When the models were re-run for site-specific cancers, all three models for DD (Figure 3) and AD (Figure 4) were positively associated with lung, blood, and liver cancers, while models for AD were also associated with prostate cancer. Contrastingly, DD and AD were associated with a reduced risk of breast cancer, showing 18% and 15% reduction for model 3, respectively. The association for BD, SZ, and PTSD could not be modelled due to insufficient statistical power.

**Figure 3.**
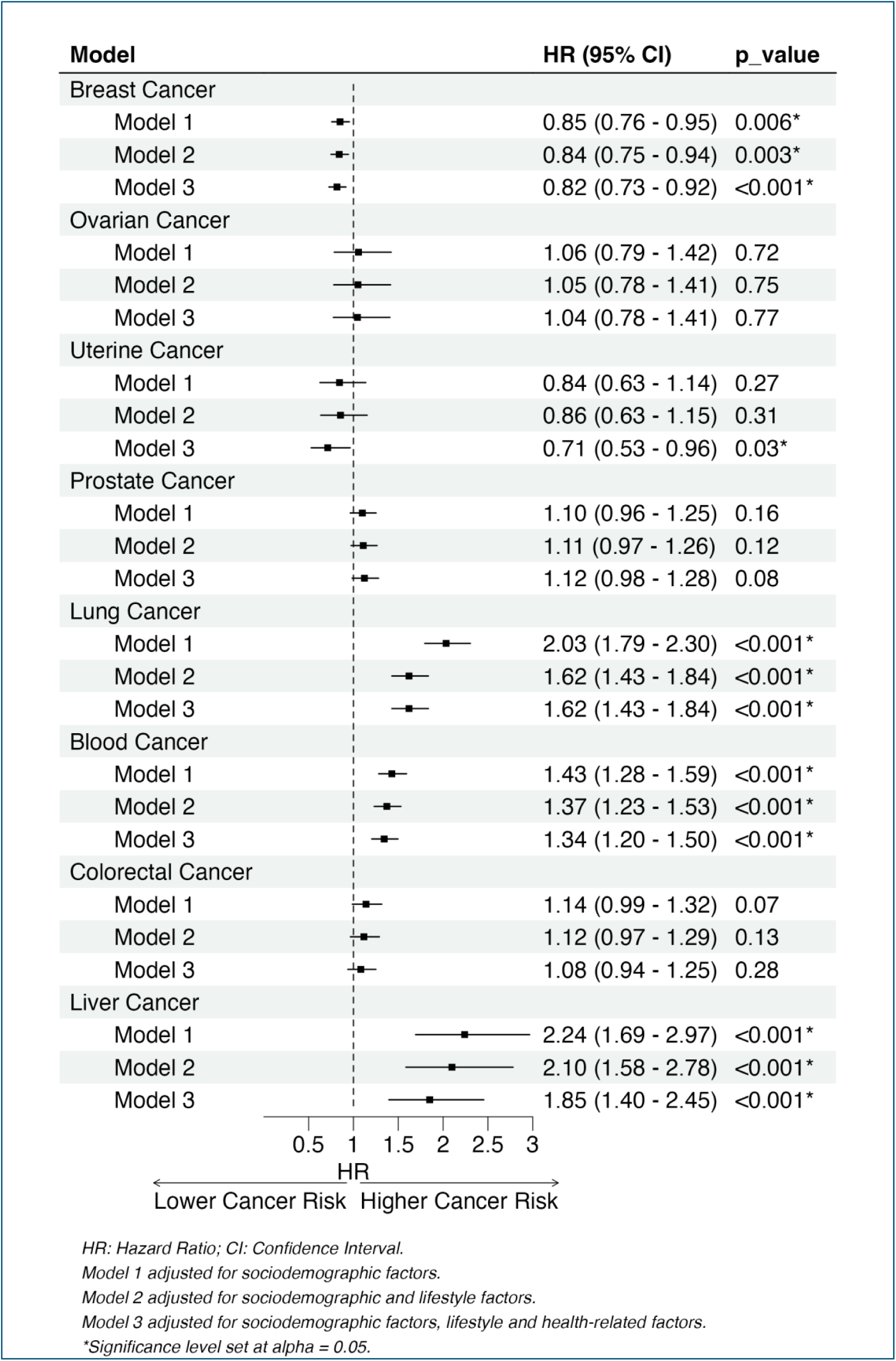
Forest Plot for Cox Models of Depressive Disorders and Site-Specific Cancer Incidence.

**Figure 4.**
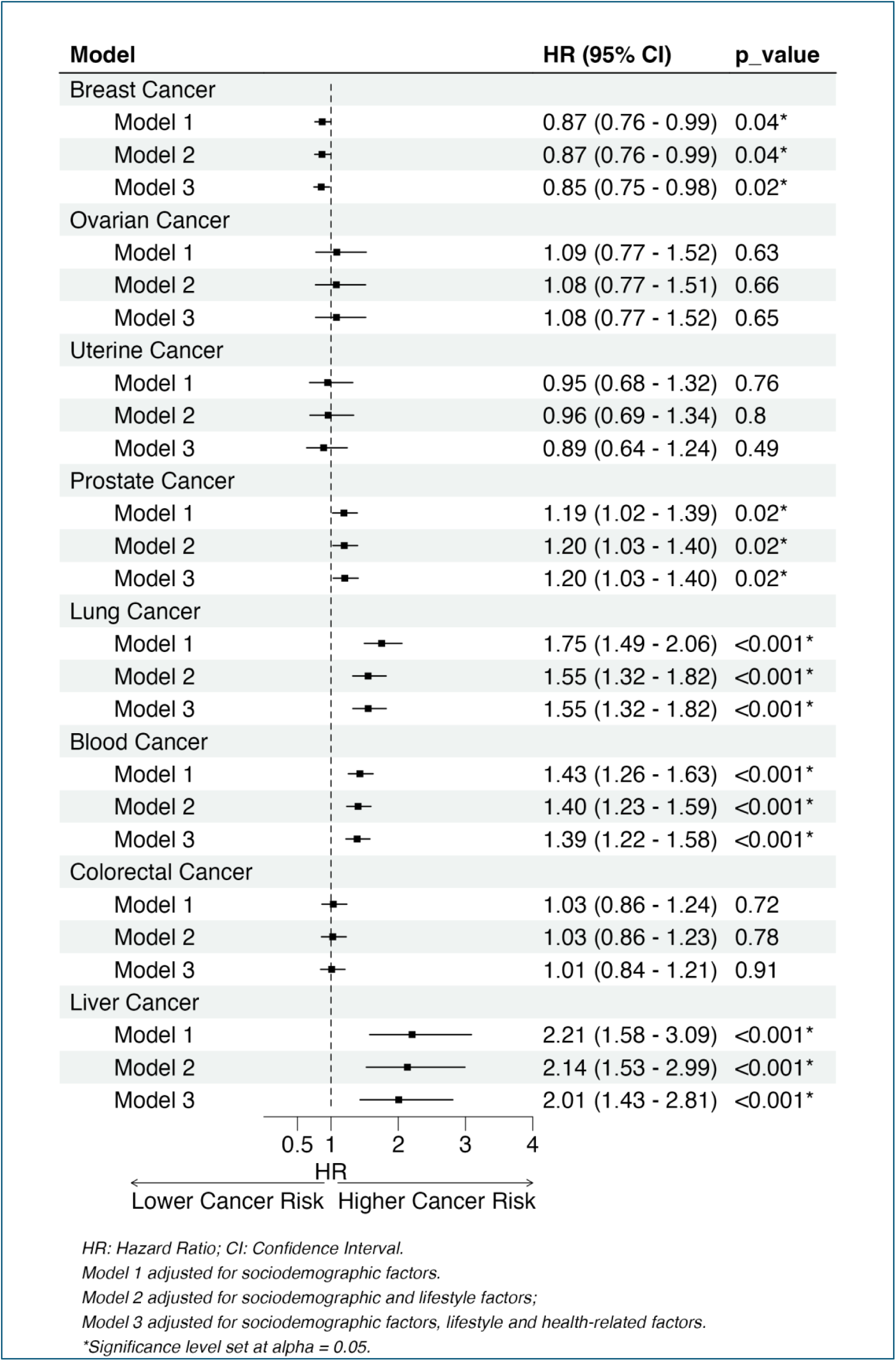
Forest Plot of Cox Models for Anxiety Disorders and Site-Specific Cancer Incidence.

After excluding 36,465 individuals with missing PHQ-4 scores, the sensitivity analysis was conducted on 365,790 participants. A positive dose-response relationship was observed between PHQ-4 score and overall cancer risk [HR: 1.03 (95%CI: 1.02-1.04)] as well as prostate, lung, blood, and liver cancer risk (Supplementary Figure 2).

## 5 Discussion

This largescale prospective population cohort study demonstrated significant associations between DD, AD, and BD and the overall risk of subsequently being diagnosed with cancer, as well as the risk of a number of site-specific cancers. These associations were independent of sociodemographic and lifestyle confounders. Interestingly, a protective association was observed for breast cancer in women, which requires further investigation. The lack of significant associations with SZ and PTSD should be corroborated in other studies or meta-analyses due to their lower statistical power.

The number of MHCs and outcomes implicated suggests possible common pathways operating across a range of conditions. Prior research supports this notion, highlighting a potential genetic overlap between MHCs [34,35], with a Brazilian study [36] identifying a 66% genetic overlap between depression and anxiety symptoms. Our hypothesis is that this shared genetic component may also contribute to increased cancer risk, possibly through pleiotropic effects; however, this remains speculative and requires further investigation in future studies.

Sex-stratified analyses revealed a male-specific association for DD and BD with overall cancer risk. This inequality likely reflects sex-based behavioural differences as men tend to engage more frequently in behaviours that are recognised risk factors for both MHCs and cancer, such as smoking and alcohol consumption. Even though we adjusted for these factors, there could be residual confounding. The UK’s Office for National Statistics 2022 report [37] and the 2021 Health Survey for England [38] highlight these differences, showing higher smoking (14.6%) and alcohol consumption (32%) prevalence among men compared to women (11.2%, 12%, respectively). Additionally, lower healthcare-seeking behaviour among men with MHCs [39] may have also contributed to this inequality. In the UK, between 2020 and 2021, the number of men (n=469,756) referred to the Improving Access to Psychological Therapies (IAPT) programme, designed to address depression and anxiety [40], was almost half that of women (n=975,399) [41]. This underrepresentation could lead to delayed diagnoses and potentially more advanced disease stages in men, influencing the observed association with overall cancer risk.

Regarding site-specific cancer risk, DD and AD were associated with higher risks of several cancers, yet displayed a potential protective effect against breast cancer. Our findings are consistent with previous studies [10,42] and may reflect differential uptake of breast cancer screening programmes rather than an actual protective effect. Since 1988, UK women aged 50-70 years have been invited to the Breast Cancer Screening Programme (BCSP) every three years [43]. However, the programme relies on voluntary participation, possibly leading to inequalities in breast cancer detection rates. A study [44] analysing BCSP uptake between 2011 and 2014 revealed 15% lower odds of attending screenings for women with MHCs. Given our sample’s eligibility for screening, this lower participation rate might explain the observed lower breast cancer risk associated with MHC diagnosis in our group. That is, the apparent lower risk could partially reflect a lower detection rate, which warrants future studies. If this mechanism is true, then people with MHCs should be diagnosed with breast cancer at a later stage. Therefore, future research is required to compare breast cancer stage at diagnosis in people with and without MHCs, as well as longer follow-up to account for possible later detection among people with MHCs.

Previous research predominantly focused on the association between depression or anxiety and overall cancer risk [12,14,15,24,45], likely reflecting their higher prevalence worldwide [46]. A consistent pattern emerged among most studies demonstrating an elevated cancer risk among individuals with these conditions [10,12,15,17]. Nonetheless, a UK investigation based on the Whitehall II study [45] reported no significant association between depression and overall cancer risk. Although this finding seemingly contradicts our observation, several methodological limitations could have impacted their findings. The study utilised historical NHS records from 1991 to detect cancer incidence, which were recognised as very incomplete; resulting in 34% missing data in the study [47]. Additionally, the study had much lower statistical power than ours, with a sample size of 6,983 and 776 incident cancer cases, compared with 402,255 and 68,065, respectively, in our study.

Few studies have examined the associations with different cancer sites, with lung cancer being the most studied. The current study aligns with prior evidence showing a higher risk of lung cancer associated with depression [11,24] and anxiety [48]. Evidence linking MHCs to other cancers is even scarcer. To our knowledge, only one study [45] investigated the association between depression and liver cancer risk and reported no association (OR: 0.81, 95%CI: 0.59-1.11). However, this finding was based on a retrospective investigation of 114 participants. Hence, further research is needed to replicate our findings.

The mechanisms linking MHCs to cancer risk are unclear, but our findings suggest that lifestyle factors may play a mediating role in some cancers. In general, adjustment for lifestyle factors attenuated associations slightly, but they remained significant. For the associations between DD and cancer sites, attenuation following adjustment for lifestyle was most apparent for lung cancer, followed by liver cancer, suggesting these associations may be mediated in part by lifestyle. Smoking has previously been postulated as a mediator of the association between depression and lung cancer [11], and alcohol consumption, a recognised risk factor for liver cancer [49], can increase as a result of depression [50].

Our study had a number of strengths. First, the large UK Biobank sample ensured sufficient power to examine associations across five MHCs and eight cancer sites, as well as testing of interactions with sex and subgroup analyses. Second, utilising administrative data to ascertain outcomes minimised the risk of information bias. Third, the study involved a comprehensive set of covariates, allowing for the adjustment of most recognised confounders. Last, the study’s long follow-up provided a sufficient observation period for cancer development, as a previous systematic review [51] showed that studies with less than 10-year follow-up were less likely to detect significant associations.

However, the study has limitations. As an observational study, it cannot establish causation. Also, despite thorough covariate adjustment, residual or unmeasured confounding cannot be ruled out. For instance, previous research suggests a potential influence of antidepressant use on cancer risk [52,53]. However, this data is not well-captured in the UKB.

MHC diagnoses in our study were not mutually exclusive. Participants were classified as having a given condition if they had a recorded diagnosis for that disorder, regardless of co-occurrence with other conditions. This may have inflated the estimated risk for individuals with multiple co-occurring conditions. As such, these estimates likely reflect the cumulative burden of mental health comorbidity rather than isolated effects.

Furthermore, the prevalence of MHCs at baseline included self-report of physician diagnoses, but MHCs diagnosed thereafter were ascertained from hospital records, potentially excluding less severe cases managed exclusively in the community [54]. Nonetheless, the observed median PHQ-4 score of 2 within the sample suggests that the analysis captured a range of MHC severities, including less severe cases. The reliance on hospital admissions or deaths to ascertain cancer outcomes may have resulted in incomplete ascertainment of early and less severe cases managed in the community or outpatients; however, this would mainly apply to skin cancers, which were not included in the study.

The PH assumption violations in model 3 of DD-overall cancer, DD-breast cancer, AD-breast, AD-ovarian, and AD-colorectal cancer may affect HR interpretation [55]. For these models, HRs should be interpreted as a weighted average effect over the study period, as suggested in a recent study [56]. Lastly, the UK Biobank’s low baseline response rate (5.5%) raises concerns about its representativeness. However, previous studies [57] showed that associations obtained from the UK Biobank were broadly consistent with those from more representative studies, supporting their generalizability.

In conclusion, a range of MHCs (DD, AD and BD) were associated with higher risk of cancer overall and several site-specific cancers. While causality cannot be assumed, diagnoses of MHCs could be useful for cancer risk stratification, as these individuals might represent a high-risk group. Furthermore, the findings reinforce the need to support healthy lifestyles among people with MHCs.

## Data Availability

The data used in this study are available from UK Biobank (www.ukbiobank.ac.uk) upon application. Access to the data requires approval from the UK Biobank and is subject to their data access policies. The authors do not have permission to share the data directly.

## 6 Acknowledgements

We thank all UK Biobank participants. This study was conducted under the project 71392.

## 7 Authors’ Contributions

MSA contributed to the study conception, conducted the analyses, interpreted the data and drafted the manuscript. FH proposed the idea, contributed to the analysis, and contributed to drafting the manuscript. SPS, ZZ, SN, IDR, CCM, and JPP critically reviewed the manuscript and approved the final draft. FH and MSA are the guarantors of this work and, as such, had full access to all the data in the study and take responsibility for the integrity of the data and the accuracy of the data analysis.

