## Supplementary Table 1 for "Mental health conditions and incident cancer: a prospective cohort study of 402,255 UK Biobank participants"

Supplementary Table 1. Summary Statistics of Cancer Incidence Stratified by MHCs diagnosis

| Cancer Incidence[1] | **All MHCs** N = 39,304 | **DD** N = 25,616 | **AD** N = 21,892 | **BD** N = 1,510 | **SZ** N = 1,719 | **PTSD** N = 477 |
|---|---|---|---|---|---|---|
| Overall | 9,122 (23%) | 5,839 (23%) | 5,303 (24%) | 309 (20%) | 358 (21%) | 86 (18%) |
| Site-specific Cancer | | | | | | |
| Breast | 1,679 (4.3%) | 1,043 (4.1%) | 1,061 (4.8%) | 51 (3.4%) | 39 (2.3%) | 15 (3.1%) |
| Ovarian | 239 (0.6%) | 150 (0.6%) | 139 (0.6%) | 10 (0.7%) | 11 (0.6%) | 3 (0.6%) |
| Uterine | 271 (0.7%) | 169 (0.7%) | 169 (0.8%) | 10 (0.7%) | 7 (0.4%) | 1 (0.2%) |
| Prostate | 1,143 (2.9%) | 739 (2.9%) | 615 (2.8%) | 53 (3.5%) | 51 (3.0%) | 15 (3.1%) |
| Lung | 874 (2.2%) | 579 (2.3%) | 481 (2.2%) | 30 (2.0%) | 44 (2.6%) | 5 (1.0%) |
| Blood | 1,463 (3.7%) | 907 (3.5%) | 859 (3.9%) | 49 (3.2%) | 69 (4.0%) | 14 (2.9%) |
| Colorectal | 873 (2.2%) | 547 (2.1%) | 516 (2.4%) | 27 (1.8%) | 36 (2.1%) | 10 (2.1%) |
| Liver | 144 (0.4%) | 96 (0.4%) | 75 (0.3%) | 4 (0.3%) | 13 (0.8%) | 2 (0.4%) |

[1]n (%); MHCs: mental health conditions; DD: depressive disorders; AD: anxiety disorders; BD: bipolar disorders; SZ; schizophrenia; PTSD: post-traumatic stress disorders
