## Supplementary Table 2 for "Mental health conditions and incident cancer: a prospective cohort study of 402,255 UK Biobank participants"

Supplementary Table 2. Baseline Characteristics of Included Participants Stratified by Sex (n = 402,255)

| Characteristic | Overall<br>N = 402,255 | Female<br>N = 214,015 | Male<br>N = 188,240 | p-value[4] |
|---|---|---|---|---|
| Age[1] (years) | 56, (8) | 56, (8) | 57, (8) | <0.001 |
| Ethnicity[2] | | | | <0.001 |
| White | 380,847 (95%) | 202,326 (95%) | 178,521 (95%) | |
| Black | 6,235 (1.6%) | 3,624 (1.7%) | 2,611 (1.4%) | |
| Chinese | 1,277 (0.3%) | 810 (0.4%) | 467 (0.2%) | |
| Mixed | 2,360 (0.6%) | 1,470 (0.7%) | 890 (0.5%) | |
| South Asian | 7,975 (2.0%) | 3,764 (1.8%) | 4,211 (2.2%) | |
| Any other | 3,561 (0.9%) | 2,021 (0.9%) | 1,540 (0.8%) | |
| Deprivation[1] | -1.37 (3.04) | -1.40 (2.99) | -1.35 (3.09) | <0.001 |
| Smoking Status[2] | | | | <0.001 |
| Current | 41,452 (10%) | 18,601 (8.7%) | 22,851 (12%) | |
| Never | 219,829 (55%) | 127,444 (60%) | 92,385 (49%) | |
| Previous | 140,974 (35%) | 67,970 (32%) | 73,004 (39%) | |
| Alcohol[3] (units/week) | 11 (3, 23) | 8 (2, 15) | 18 (8, 33) | <0.001 |
| Sleep Duration[1] (hour) | 7.16 (1.08) | 7.18 (1.09) | 7.13 (1.06) | <0.001 |
| Total physical activity (MET-min/wk)[1] | 2,422 (2,439) | 2,267 (2,282) | 2,599 (2,595) | <0.001 |
| Processed Meat Consumption[1] | | | | <0.001 |
| Never | 37,547 (9.3%) | 27,319 (13%) | 10,228 (5.4%) | |
| Less than once a week | 121,657 (30%) | 81,519 (38%) | 40,138 (21%) | |
| Once a week | 117,335 (29%) | 61,335 (29%) | 56,000 (30%) | |
| 2-5 times a week | 109,667 (27%) | 39,962 (19%) | 69,705 (37%) | |
| 5-6 times a week | 12,787 (3.2%) | 3,102 (1.4%) | 9,685 (5.1%) | |
| once or more daily | 3,262 (0.8%) | 778 (0.4%) | 2,484 (1.3%) | |
| BMI[1] (kg/m$^2$) | 27.3 (4.7) | 26.9 (5.1) | 27.8 (4.2) | <0.001 |
| Self-reported Diabetes[2] | 18,006 (4.5%) | 6,483 (3.0%) | 11,523 (6.1%) | <0.001 |
| SBP (mmHg) | 138 (19) | 135 (19) | 141 (17) | <0.001 |
| Menopausal Status[2] | | | | <0.001 |
| no | 52,377 (13%) | 52,377 (24%) | 0 (0%) | |
| yes | 128,630 (32%) | 128,630 (60%) | 0 (0%) | |
| Not applicable | 221,248 (55%) | 33,008 (15%) | 188,240 (100%) | |
| Follow-up Time[3] (year) | 13.4 (12.6, 14.3) | 13.5 (12.7, 14.3) | 13.4 (12.5, 14.2) | <0.001 |

[1]Mean, (SD); [2]n (%); [3]Median (IQR)
[4]Welch Two Sample t-test; Pearson's Chi-squared test; Wilcoxon rank sum test
Significance level set at alpha = 0.05
MET: Metabolic Equivalent Task; BMI: Body Mass Index; SBP: Systolic Blood Pressure
