## Supplementary Table 3 for "Mental health conditions and incident cancer: a prospective cohort study of 402,255 UK Biobank participants"

### Supplementary Table 1. Likelihood Ratio Tests Comparing Cox Models With and Without Interaction Terms Between MHCs and Sex in Relation to Cancer Incidence

| Interaction Term | LRT[1] | df | P-value |
|---|---|---|---|
| Depression | 6.021 | 1 | 0.0141* |
| Anxiety | 2.319 | 1 | 0.1278 |
| Bipolar disorder | 5.836 | 1 | 0.0157* |
| SZ | 1.203 | 1 | 0.2728 |
| PTS | 0.005 | 1 | 0.9445 |

*[1]Likelihood Ratio Tests*

*Each comparison tests whether including an interaction term between the specified mental health condition and sex significantly improves model fit. All models are adjusted for age, ethnicity, deprivation, smoking status, alcohol intake, physical activity, sleep duration, processed meat intake, BMI, diabetes, and systolic blood pressure.*
