## Supplementary Figure 1 for "Mental health conditions and incident cancer: a prospective cohort study of 402,255 UK Biobank participants"

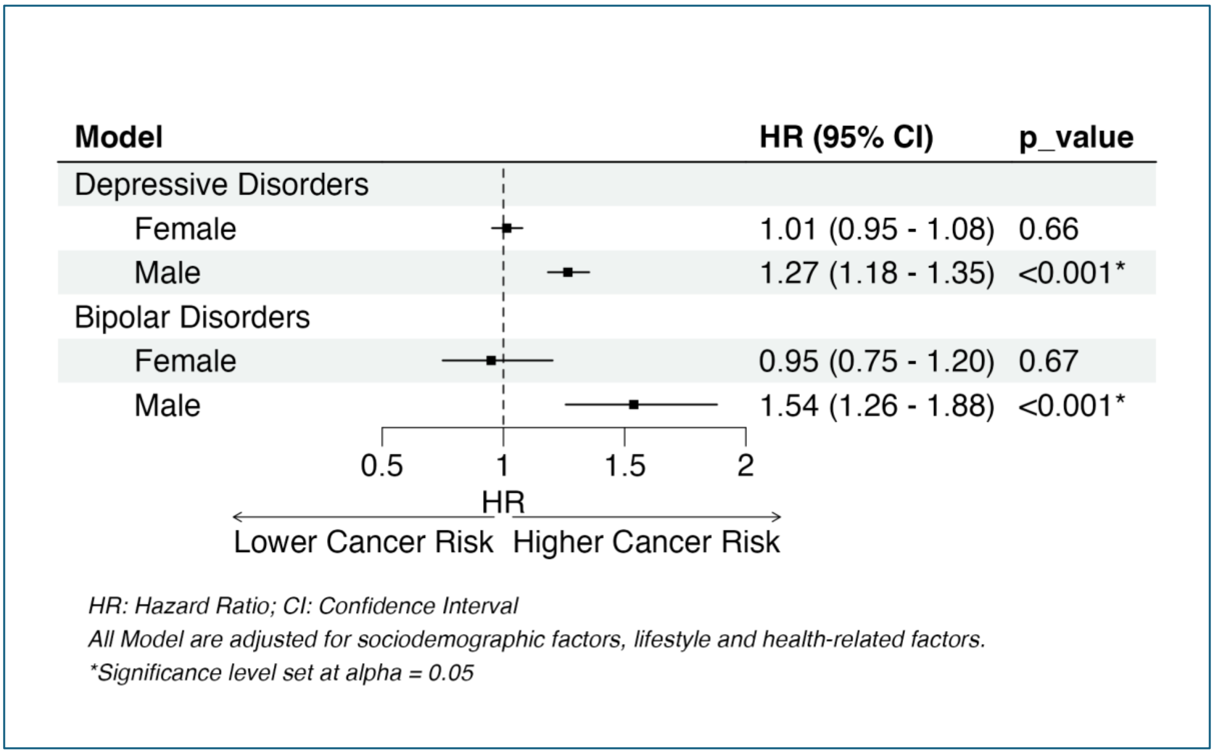


Supplementary Figure 1. Forest Plot of Cox Models for Depressive Disorders and Bipolar Disorders and Overall Cancer Stratified by Sex
