## Supplementary figures and images for "Mental health conditions and incident cancer: a prospective cohort study of 402,255 UK Biobank participants"

### Supplementary Figure 2

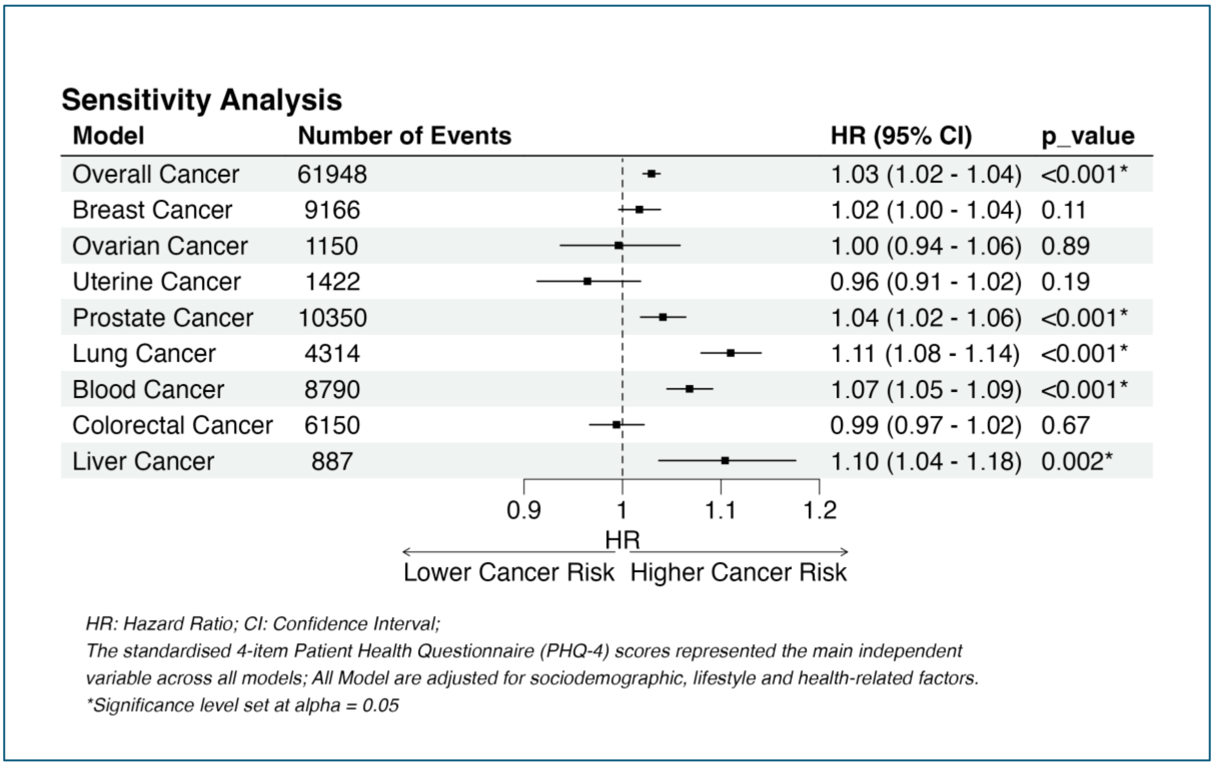


Supplementary Figure 1. Forest Plot of the Sensitivity Analysis Models using PHQ-4
